# Mental health in relation to changes in sleep, exercise, alcohol and diet during the COVID-19 pandemic: examination of four UK cohort studies

**DOI:** 10.1101/2021.03.26.21254424

**Authors:** Aase Villadsen, Praveetha Patalay, David Bann

## Abstract

**Background:** Responses to the COVID-19 pandemic have included lockdowns and social distancing with considerable disruptions to people’s lives. These changes may have particularly impacted on those with mental health problems, leading to a worsening of inequalities in the behaviours which influence health.

**Methods:** We used data from four national longitudinal British cohort studies (N=10,666). Respondents reported mental health (psychological distress and anxiety/depression symptoms) and health behaviours (alcohol, diet, physical activity, and sleep) before and during the pandemic. Associations between pre-pandemic mental ill-health and pandemic mental ill-health and health behaviours were examined using logistic regression; pooled effects were estimated using meta-analysis.

**Results:** Worse mental health was related to adverse health behaviours; effect sizes were largest for sleep, exercise and diet, and weaker for alcohol. The associations between poor mental health and adverse health behaviours were larger during the May lockdown than pre-pandemic. In September, when restrictions had eased, inequalities had largely reverted to pre-pandemic levels. A notable exception was for sleep, where differences by mental health status remained high. Risk differences for adverse sleep for those with the highest level of prior mental ill-health compared to those with the lowest, were 21.2% (95% CI: 16.2, 26.2) before lockdown, 25.5% (20.0, 30.3) in May, and 28.2% (21.2, 35.2) in September.

**Conclusions:** Taken together, our findings suggest that mental health is an increasingly important factor in health behaviour inequality in the COVID era. The promotion of mental health may thus be an important component of improving post-COVID population health.

## Introduction

Health behaviours such as exercise, sleep, diet, alcohol use, and smoking are important modifiable contributors to the global burden of disease—such as diabetes, heart disease, and cancer (Khaw et al., 2008). Furthermore, health behaviours have been linked to mental health and wellbeing, with studies demonstrating that those with mental health problems are more likely to engage in unhealthy behaviours (Jane-Llopis & Matytsina, 2006; Lasser et al., 2000; Stranges, Samaraweera, Taggart, Kandala, & Stewart-Brown, 2014). The COVID-19 pandemic and associated lockdown and home confinement is likely to have had an impact on health behaviours as this new way of life may have led to changes in exercise regimes, dietary and sleeping patterns, and alcohol and tobacco use (Ammar et al., 2020; Biddle, Edwards, Gray, & Sollis, 2020; Cellini, Canale, Mioni, & Costa, 2020; Deschasaux-Tanguy et al., 2020; Di Renzo et al., 2020; Duffy, 2020; Wardell et al., 2020). Previous research has highlighted socio-demographic inequalities in changes in health behaviours during the pandemic (Bann et al., 2020; Biddle et al., 2020; Deschasaux-Tanguy et al., 2020; Giustino et al., 2020; Koopmann, Georgiadou, Kiefer, & Hillemacher, 2020). However, such behaviours may also differ as a result of individual level health factors, such as mental health status (Stanton et al., 2020); these links may in turn lead to a worsening of subsequent mental and physical health outcomes.

It is conceivable that those with poor mental health may be especially susceptible to detrimental lifestyle changes during the pandemic. Existing studies have examined inequalities in health behaviours based on mental health. These are largely cross-sectional in nature, and have suggested that poor mental health is detrimental to some health behaviours during the pandemic (Cellini et al., 2020; Cheval et al., 2020; Deschasaux-Tanguy et al., 2020; Stanton et al., 2020; Xiao, Zhang, Kong, Li, & Yang, 2020). However, previous studies have been limited in terms of sample representativeness. Moreover, previous studies have been limited to examining mental health concurrent with the pandemic rather than considering mental health status prior to this event.

The current study addresses this gap by examining mental health prior to the pandemic as a predictor of health behaviour immediately before and at two timepoints during the pandemic. This enables comparisons of associations during the height of the first UK lockdown (May 2020) and later in the pandemic when some restrictions had eased (September 2020). We were thus able to investigate if the pandemic led to a widening of such inequalities in health behaviours by mental health status. We used data from four nationally representative UK cohort studies, representing different age groups (19-20, 30-31, 50, and 62 years). Measures of mental health were also obtained from the previous survey sweep in each of the respective cohort studies. Since the magnitude of association and its change across the course of the pandemic may differ by age and sex, we formally tested for heterogeneity by cohort and sex (Alati et al., 2004; Gibson, 2012).

## Methods

### Sample

Data are from four UK longitudinal cohort studies. The National Child Development Study (NCDS) is the oldest cohort, follows the lives of an initial 17,415 people born in 1958 (Power & Elliott, 2006). The 1970 British Cohort Study (BCS70) is based on initially 17,196 cohort members born in 1970 (Elliott & Shepherd, 2006). The Next Steps cohort are born in 1989 starting with 15,770 cohort members (Calderwood & Sanchez, 2016). Finally, the youngest cohort, the Millennium Cohort Study (MCS), began with an original sample of 18,818 born in 2001 (Joshi & Fitzsimons, 2016). In this paper we refer to these cohorts according to the year participants were born, so 1958c, 1970c, 1989c, and 2001c. The cohorts have been followed up at regular intervals from birth, with exception of the 1989 cohort which was recruited at age 14. Measures and assessments have been broad, spanning across the domains of health, mental health, socioeconomics, and demographics. All cohorts were administered an online questionnaire during the height of the COVID-19 pandemic lockdown in May 2020, and again in September 2020 when some restrictions had eased. The COVID-19 survey was issued to a sample of nearly 39,000 across the four cohorts for whom an email address was held and who had not attritted permanently from their respective cohort study. Nearly 14,000 responded to the first survey in May that captured various aspects of their lives during the pandemic, including health behaviours. Analyses in the current study are based on 10,666 participants who provided valid responses to questions on health behaviours before and during the pandemic in the May survey and again in the September survey. Further information on the COVID-19 survey is available elsewhere (Brown et al., 2020).

## Measures

### Health behaviour

Four aspects of health behaviour outcomes were measured (alcohol, diet, exercise, and sleep). In the first survey in May 2020 participants reported their behaviours in the month before the Coronavirus outbreak and their current behaviours, and the second survey in September 2020 again asked about current behaviours. For each health behaviour, binary measures were constructed distinguishing healthy and risky behaviour using recommended guidelines. Main conclusions did not differ when using all groups (data available upon request). Alcohol consumption was measured in terms of frequency (frequency from never to 4 or more time a week) and volume (number of drinks per typical day when drinking). From this, a measure of risky drinking was constructed using current UK guidelines recommending no more than 14 units a week (National Health Service, 2018b), and less than six units in a session (National Health Service, 2019a). Because our survey asked about drinks rather units, our thresholds were up to 12 drinks weekly and less than five drinks per session. Diet was ascertained in number of portions of fresh fruit and vegetables consumed in a typical day, from which a binary measure was created using the ‘five a day’ recommendation as a cut off (National Health Service, 2018a). Physical activity was measured as number of days per week doing exercise for at least 30 minutes that raises the heart rate and causes sweating; a binary measure was constructed with a cut-off point of less than 5 days a week falling short of the recommended 150 minutes a week (National Health Service, 2019b). Finally, sleep was reported as average hours per night, which was dichotomised into a variable distinguishing recommended sleep levels (7-9 hours) versus atypical sleep (<7 or >9) (Hirshkowitz et al., 2015).

### Mental health

Multiple psychological health measures were used: 1) psychological distress (measured using different scales in each cohort, both prospectively before COVID-19 and during first lockdown) and 2) anxiety and depression symptoms (ascertained during lockdown in May using the same scale across the cohorts). Each has complementary advantages—the former in mapping hypothesised temporal directions using well characterised measures used longitudinally in each cohort and the latter in terms of improving comparability for testing cohort differences in association; thus both were used separately in analyses.

Psychological distress prior to the pandemic was measured using different scales in each cohort. In the 2001c this was at age 17 (two years prior) using the Kessler (K6), a six item measure ranging 0-24, with scores of 13 and above considered in the clinical range (Kessler et al., 2003). In the 1989c the assessment was at age 25 (five years prior), using the General Health Questionnaire (GHQ-12), ranging from 0-12, with clinical level of 4 and above (Goldberg & Williams, 1988). In the 1970c the assessment was at age 46 (four years prior), and in the 1958c at age 50 (12 years prior), both using the 9-item Malaise, ranging from 0-9 with scores of 4 or above considered in the clinical range (Rutter, Tizard, & Whitmore, 1970). These cohort specific measures were administered also in the COVID-19 survey in May and are referred to in this study as current psychological distress. High psychological distress in the current study is the established clinical cut off for each of these respective measures.

Assessments in the cohorts since the start of the COVID-19 pandemic include: depressive symptoms were measured using two items from the Patient Health Questionnaire (PHQ-2), range 0-6, and scores of 3 and above is indicative of high depressive symptoms (Kroenke, Spitzer, & Williams, 2003). Anxiety symptoms were assessed by two items from the General Anxiety Disorder scale (GAD-2), range 0-6, with scores of 3 and above considered high levels of anxiety symptoms (Kroenke, Spitzer, Williams, Monahan, & Lowe, 2007). These scales were combined into one single measure of anxiety/depression, range 0-12, and high levels of symptoms were set to 6 or above, a threshold that was guided by the distribution of cut-offs for the two subscales.

### Education attainment

Since education may influence both mental health and health behaviours (Huijts et al., 2017; Yu & Williams, 1999), it was included as a potential confounder. Cohort members’ level of education was classified using the National Vocational Qualifications (NVQ) level system, ranging from NVQ1 to NVQ5, with an additional category for those without any qualifications. For the youngest cohort, parental educational level was used as many were still in training or education.

## Analyses

All statistical analyses were carried out using Stata version 16 (StataCorp, 2019). We examined how prior mental health (psychological distress) and mental health during the lockdown in May (psychological distress, and anxiety/depression), was associated with health behaviour at three timepoints: the month before the Coronavirus outbreak, during the lockdown in May, and in September when restrictions had eased. Descriptive statistics and unadjusted associations between mental health and health behaviour used clinical cut-offs (binary measures) of mental health. In logistic regression models, adjusting for gender and for educational level of cohort members, ridit scores of mental health were used to estimate inequalities in each behaviour, to maximise statistical power and avoid information loss. When used in regression models, it is referred to as the slope index of inequality and provides a single estimate of the total magnitude of association (inequality), while accounting for differences in the distribution of participants within each cohort (World Health Organzation, 2017). Where the prevalence of the outcome differs across time, comparing results on the relative scale can impair comparisons of risk factor-outcome associations (e.g., identical odds ratios can reflect different associations on the absolute scale) (King, Harper, & Young, 2012). As such, absolute risk differences in health behaviour outcomes by mental health were obtained using the margins command in Stata following logistic regression. Effect estimates show the difference in risk for each outcome comparing those with the highest compared with least mental health symptoms.

Regression analyses were carried out by cohort and results were meta-analysed to formally assess heterogeneity using the I^2^ statistic, and to obtain the overall pooled association between health behaviour risk and mental health. Models examining cohort estimates controlled for gender and education. In the models examining gender differences, educational level and cohort were controlled for. Additional analyses were carried out to check if regression results were consistent when using continuous standardised and binary measures of mental health.

In all analyses, bias due to non-response to the survey was adjusted for by using weights. These were generated in logistic regression models where the outcome was response to the COVID-19 survey, and predictors were a number of demographic, socioeconomic, household and individual level variables (Brown et al., 2020). We also accounted for the stratified survey designs of the 1990c and 2001c in all analyses.

### Additional and sensitivity analyses

To examine if the conclusions differed in each gender, we conducted additional gender-specific analyses. We also conducted analyses using mental health exposures as continuous standardised scores and as binary variables instead of ridit scores.

## Results

Shown in Table 1 are sample characteristics by mental health status. For current anxiety/depression, in which the same questions were asked across all cohorts, symptoms were considerably more prevalent in younger cohorts (e.g. 26.7% [CI: 23.0, 30.8] in 2001c, and 17.5% [CI: 14.4, 21.0] in 1989c, compared with 9.2% [CI: 7.5-11.4] in 1970c and 7.8% [6.5-9.2] in 1958c). Similar patterns were found for both prior and current psychological distress (using cohort specific measures). Lower education was related to worse mental health, with associations being most consistent for psychological distress. For all mental health measures, prevalence was higher in females than in males.

**Table 1:** Sample characteristics by mental health.

| Sample characteristics | Whole sample | Psychological distress<br>(prior to pandemic) |  | Psychological distress<br>(during May lockdown) |  | Anxiety and depression<br>(during May lockdown) |  |
| --- | --- | --- | --- | --- | --- | --- | --- |
|  |  | % above clinical threshold |  | % above clinical threshold |  | % above clinical threshold |  |
|  |  | 95% CI |  | 95% CI |  | 95% CI |  |
| Whole sample | N=10,666 | 18.5% | 17.2%, 19.8% | 18.0% | 16.8%, 19.2% | 12.4% | 11.3%, 13.6% |
| Cohorts |  |  |  |  |  |  |  |
| 2001c (age 19-20) | N=1,615 | 18.0% | 15.2%, 21.2% | 18.7% | 15.7%, 22.1% | 26.7% | 23.0%, 30.8% |
| 1989c (age 30-31) | N=1,462 | 26.7% | 23.1%, 30.8% | 35.9% | 32.0%, 39.9% | 17.5% | 14.4%, 21.0% |
| 1970c (age 50) | N=3,235 | 20.4% | 18.0%, 23.0% | 17.0% | 14.9%, 19.3% | 9.2% | 7.5%, 11.4% |
| 1958c (age 62) | N=4,354 | 14.2% | 12.5%, 16.2% | 12.7% | 11.0%, 14.5% | 7.8% | 6.5%, 9.2% |
| Gender |  |  |  |  |  |  |  |
| Females | 60.4% | 22.0% | 20.3%, 23.8% | 23.2% | 21.5%, 24.9% | 15.6% | 14.1%, 17.2% |
| Males | 39.6% | 14.5% | 12.7%, 16.5% | 12.4% | 10.8%, 14.1% | 9.0% | 7.5%, 10.8% |
| Educational level |  |  |  |  |  |  |  |
| None | 4.8% | 24.5% | 18.6%, 31.5% | 22.1% | 16.6%, 28.8% | 12.8% | 9.0%, 17.8% |
| NVQ1 level | 5.8% | 24.7% | 18.5%, 32.1% | 17.8% | 13.4%, 23.3% | 11.5% | 7.8%, 16.7% |
| NVQ 2 level | 22.8% | 18.8% | 16.7%, 21.0% | 19.1% | 17.1%, 21.2% | 14.0% | 12.0%, 16.3% |
| NVQ 3 level | 17.2% | 16.8% | 14.5%, 19.4% | 21.2% | 18.2%, 24.4% | 15.8% | 12.7%, 19.5% |
| NVQ 4 level | 40.0% | 15.5% | 13.8%, 17.4% | 15.7% | 14.1%, 17.5% | 11.1% | 9.6%, 12.9% |
| NVQ 5 level | 9.4% | 15.1% | 11.7%, 19.3% | 18.7% | 14.9%, 23.1% | 8.2% | 6.2%, 10.7% |

Note: Psychological distress prior to pandemic was measured in the 2001c at age 17, in 1989c at age 25, in 1970c at age 46, and in 1958c at age 50.
Estimates of mental health are weighted to account for survey non-response. % above clinical threshold are based on scale specific cutoffs used for each measure that indicate probable clinical diagnosis. .

### Mental health and sleep

Table 2 shows that across the sample overall, 31.5% reported adverse sleep duration prior to the pandemic, and this increased to 35.9% during the May lockdown, and increased further to 39.8% in September. Across all periods—pre-pandemic, in May, and September—all measures of worse mental health were associated with adverse sleep (Table 2 and Figure 1 for binary mental health measures). The size of these inequalities appeared to be lowest pre-lockdown, and highest during the pandemic in May and September.

**Table 2:**
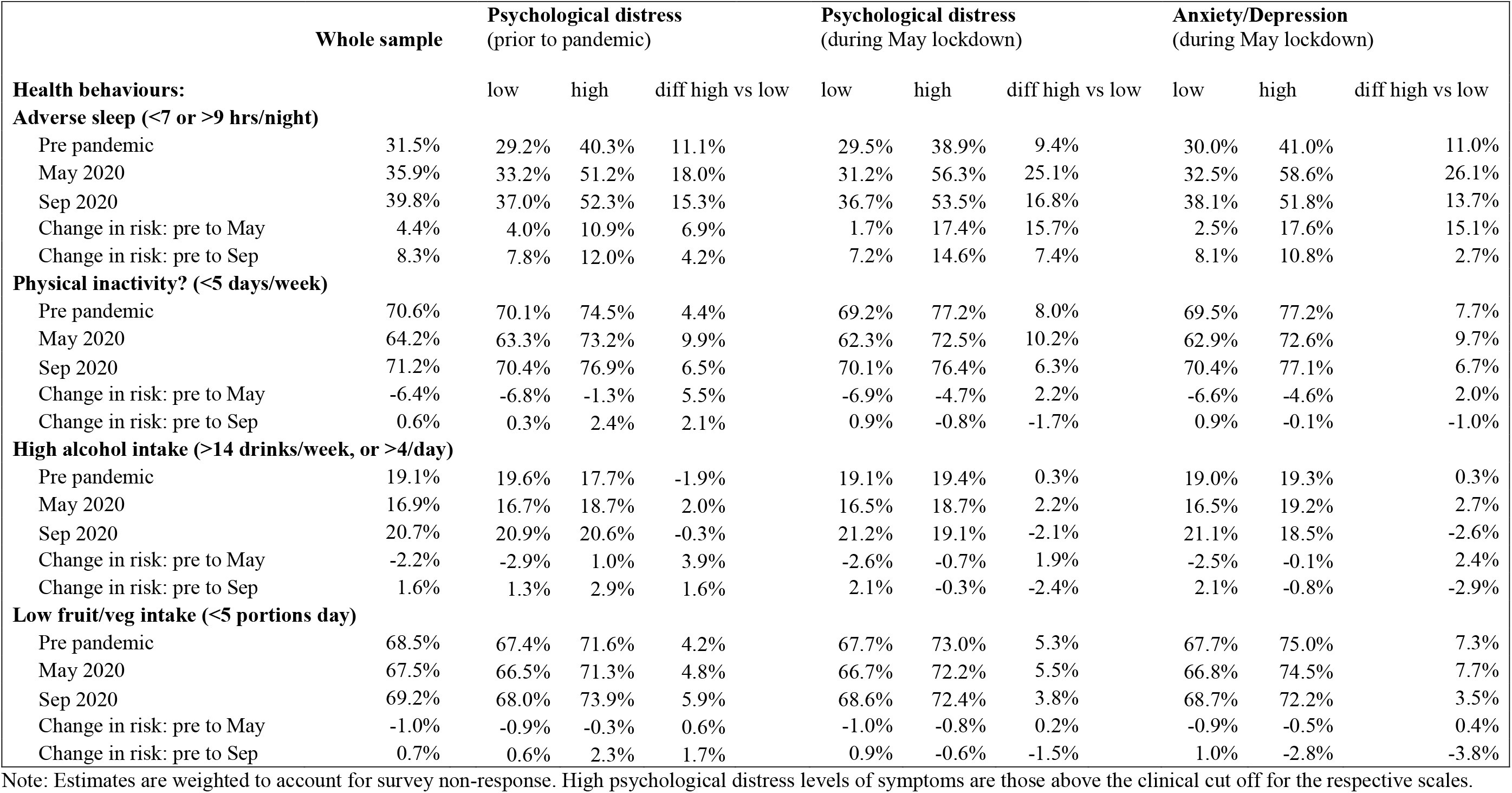
Health behaviours (before the pandemic and during May and Sept 2020) by mental health status.

**Figure 1:**
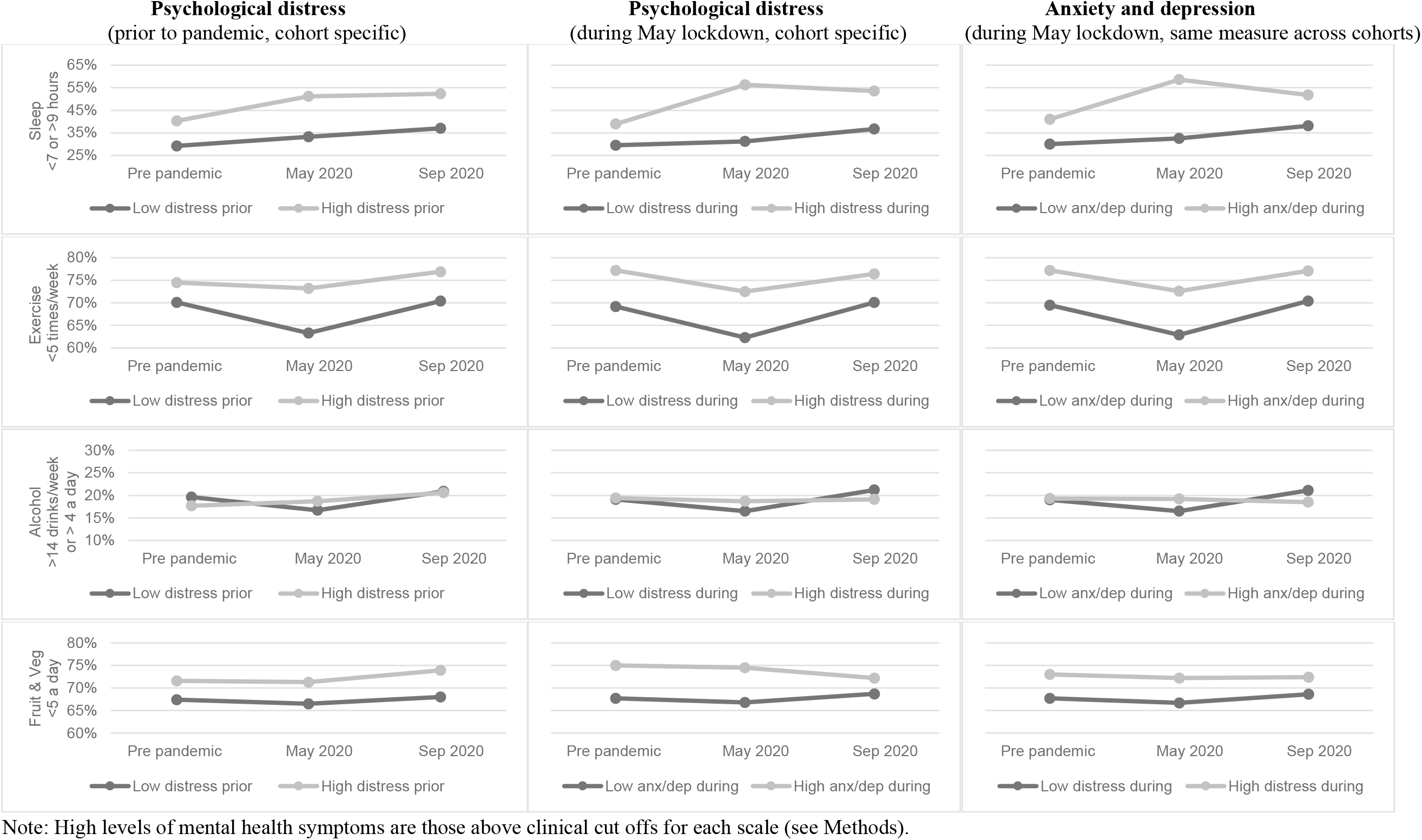
Health behaviour outcomes before and during the COVID-19 pandemic by mental health status.

The cohort-pooled risk differences for adverse sleep—in the highest compared with lowest levels of prior psychological distress—were 21.2% (95% CI: 16.2, 26.2) before lockdown, 25.5% (20.0, 30.3) in May, and 28.2% (21.2, 35.2) in September (Figure 2). There was little evidence for systematic differences by cohort (I^2^<44% in each timepoint). Findings were similar for current psychological distress and anxiety/depression (Figure 2), with effect sizes slightly weaker in September compared with May, and cohort differences were more pronounced, with the 1990c having the largest effect size in the height of the lockdown in May, yet no association prior to the pandemic.

**Figure 2.**
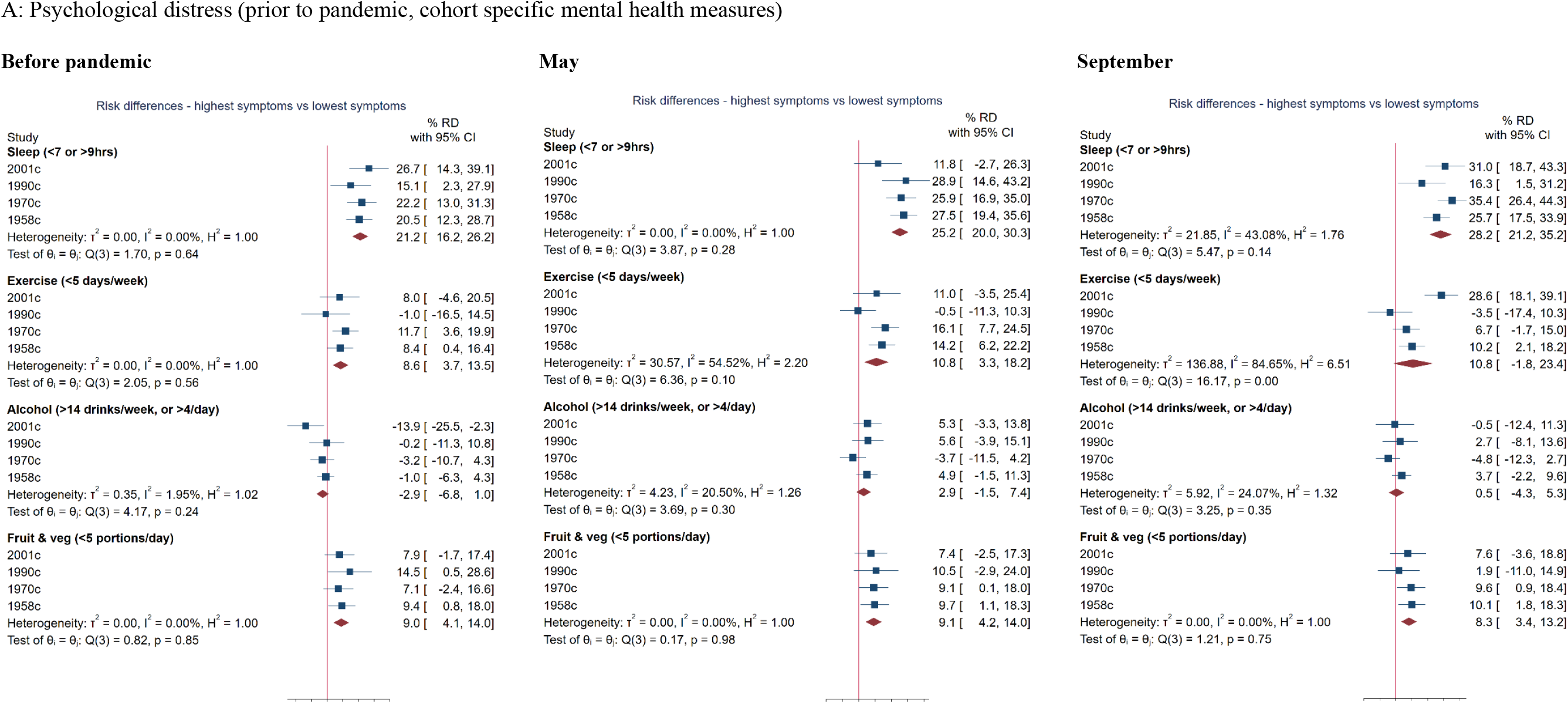
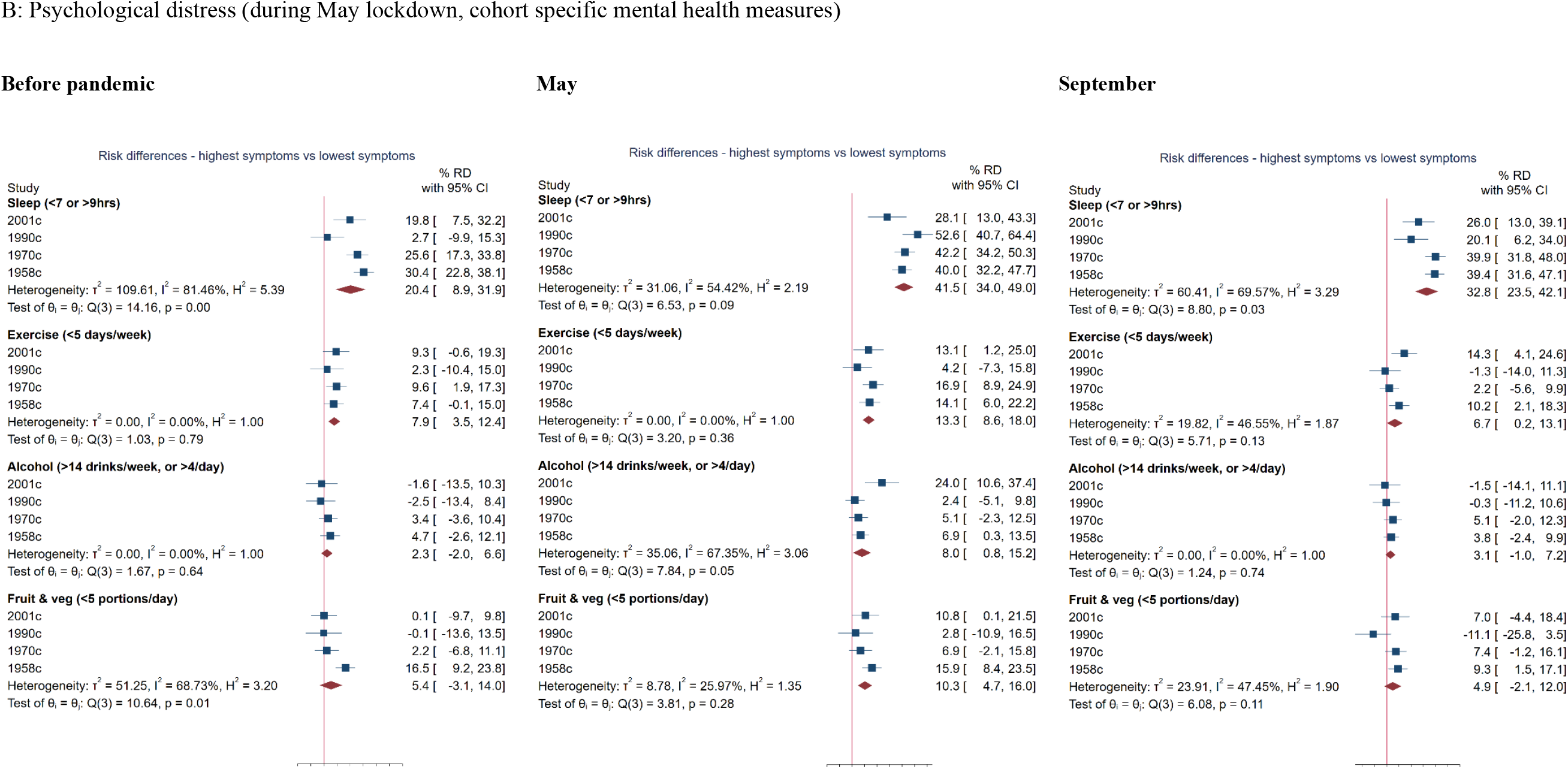
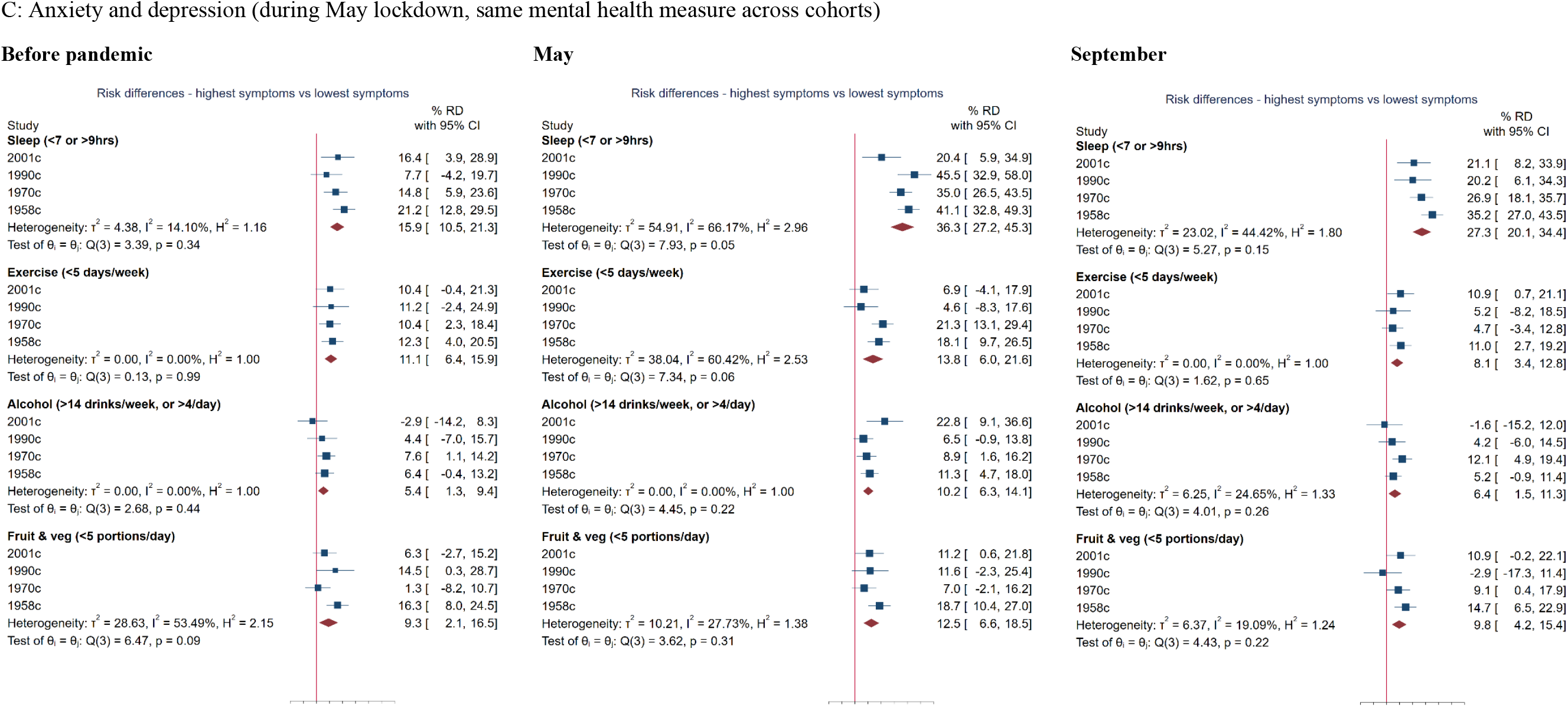
Results of logistic regressions showing differences in health behaviour risk (before COVID-19 pandemic, during May 2020 lockdown, and in September 2020) between participants with highest and lowest levels of mental health problems: meta-analysis of 5 cohort studies. Note: Estimates show the risk difference on the percentage scale between those with the highest versus lowest mental health problems (ridit scores), and are weighted to account for survey non-response and survey design in 2001c and 1990c. Sex and educational level are controlled for.

### Mental health and exercise

As shown in Table 2, prior to the pandemic 70.6% of the total sample were physically inactive during the lockdown in May this declined to 64.2%, and in September it reverted to 71.2%. Those with mental health problems (across all measures) were at greater risk of insufficient exercise before the pandemic, with inequalities increasing during the lockdown in May, and narrowing again in September (Table 2, Figure 1).

Comparing those with the highest to those with the lowest level of prior psychological distress (Figure 2), cohort-pooled risk difference for insufficient exercise were 8.5% (95% CI: 3.7, 13.5) prior to the pandemic, rising to 10.8% (95% CI: 3.3, 18.2) in May and 10.8% (95% CI: -1.8, 23.4) in September. In September inequalities were especially large in the youngest 2001c cohort (I^2^= 85%). Results for current mental health show a similar increase in inequalities from prior to the pandemic to the lockdown in May, although in September they revert to below pre-pandemic levels, and broadly there is little difference between cohorts across timepoints.

### Mental health and alcohol consumption

In terms of alcohol intake (Table 2), 19.1% reported high-risk drinking prior to the pandemic, declining to 16.9% during the lockdown in May, and then increasing to 20.7% in September. In terms of mental health and alcohol intake, the association was weak across all three timepoints (Table 2, Figure 1).

While associations between mental health and alcohol intake were largely null, for current mental health there was some evidence of inequality. For anxiety/depression the risk difference was 5.4% (95% CI: 1.3, 9.4) prior to the pandemic, rising to 10.2% (95% CI: 6.3, 14.1) in May and reverting to 6.4% (95% CI: 1.5, 11.3) in September. A similar pattern was seen for current psychological distress.

Results were largely similar across cohorts for all measures of mental health and timepoints. The only exception was for current psychological distress for which inequalities were especially large in the 2001c in May.

### Mental health and fruit and vegetable intake

As for diet (Table 2), 68.5% of the sample overall reported consuming less than five a day portions of fruit and veg before the pandemic, decreasing to 67.5% during the May lockdown and increasing to 69.2% in September. As seen in Table 2 and Figure 1, across all binary mental health measures, those with a high level of symptoms were at higher risk of not achieving the five a day recommendation at all three timepoints.

Cohort-pooled risk differences in consuming less than five a day, comparing those with the highest level of prior psychological distress to those with the lowest (Figure 2), were 9.0% (95% CI: 4.1, 14.0) prior to the pandemic, and very similar in May (9.1%, 95% CI: 4.2, 14.0) and in September (8.3%, 95% CI: 3.4, 13.2). For current mental health, we see an increase in inequalities from before to during the pandemic in May that then revert to pre-pandemic levels in September.

In term of cohort differences, for prior psychological distress results were similar across the cohorts at the three timepoints, whereas for current mental health cohorts differed before the pandemic, with inequalities greatest in the oldest 1958c cohort.

### Additional and sensitivity analyses

The main findings inequalities in health behaviours based on mental health based were similar in males and females, with few exceptions (Figure S2). Inequalities in sleep based on current mental health were greater for females than males in May. Males with a low level of prior mental health symptoms were at higher risk of excessive drinking before the pandemic, but for females there was no association. Main findings using ridit scores also did not differ when analysing mental health as either z-scores or binary variables (Figure S3).

## Discussion

### Main findings

The present study examined the association of mental health with sleep, exercise, alcohol and diet prior to and at two timepoints during the COVID-19 pandemic, using data from four UK cohort studies. For the sample overall, from before the pandemic to the full lockdown in May, there were positive improvements in exercise, diet and alcohol, but a deterioration in sleep. In September when many restrictions had eased, levels had reverted to pre-pandemic levels for most health behaviours, except for sleep for which the risk of atypical sleep had increased further.

Poor mental health was related to adverse health behaviours; especially in relation to sleep, but also exercise, and fruit and vegetable consumption, whereas for alcohol consumption the difference was small. The associations between poor mental health and health behaviour risks tended to be larger in May during the full lockdown, with 11 out of 12 effect estimates larger in May than pre-pandemic. These lockdown effects were larger for concurrently measured mental health compared to pre-pandemic measures of mental health. In September when restrictions had lessened, most health behavioural inequalities had restored to pre-pandemic levels, with 10 of 12 associations smaller in September than May. A notable exception to this general pattern of restoration was sleep, for which inequalities remained elevated into September for all measures of mental health.

### Comparison with other studies and explanations of findings

Our findings resonate well with previous research showing that poor mental health is associated with less ‘healthy’ behaviours(Jane-Llopis & Matytsina, 2006; Lasser et al., 2000; Stranges et al., 2014). Moreover, there is significant consistency between recent COVID-19 studies conducted in other countries that have examined mental health in relation to health behaviours, showing that common mental health problems such as depression and anxiety are risk factors for unfavourable changes in health behaviours during the pandemic (Cellini et al., 2020; Cheval et al., 2020; Deschasaux-Tanguy et al., 2020; Stanton et al., 2020; Xiao et al., 2020). We build on such evidence by using longitudinal nationally representative cohort data, using multiple validated mental health scales measured both prior to and during the pandemic, and also examining multiple health behavioural outcomes.

As in the current examination, an existing study has found that sleep, more so than other health behaviours, had deteriorated more during the pandemic for those with higher levels of mental health problems (Stanton et al., 2020). The strong relationship between sleep and mental health is perhaps expected as sleep is regarded as fundamental to the operation of our central nervous system and therefore linked with a large range of mental health disorders, and the relationship is highly reciprocal with mental health problems in turn being highly detrimental to sleep (Alvaro, Roberts, & Harris, 2013; Harvey, Murray, Chandler, & Soehner, 2011). This strong and cyclical relationship may explain why the sleep inequalities based on mental health had not returned to more normal pre-pandemic levels as seen for the other health behaviours. Moreover, compared to other health behaviours, sleep has a very direct or instant effect on emotional regulation (Gruber & Cassoff, 2014). In a recent review, it was proposed that the strongest pathway of the bidirectional relationship between sleep and mental health is sleep as a causal factor for the occurrence of psychiatric problems (Freeman, Sheaves, Waite, Harvey, & Harrison, 2020).

The association between mental and various other health behaviours is also likely to be reciprocal. Positive changes to health behaviours such as targeted in interventions have shown improvements in mental health following the adoption of a healthier diet (Parletta et al., 2019), smoking cessation (Taylor et al., 2014), reduced alcohol consumption (Charlet & Heinz, 2017), and increased physical activity (Atlantis, Chow, Kirby, & Singh, 2004). Conversely, the influence of mental health on subsequent health related behaviours may be the main driving mechanism for the observed higher risk of morbidity and premature mortality amongst those with mental health problems (Lawrence & Coghlan, 2002; Ploubidis, Batty, Patalay, Bann, & Goodman, 2021; Reilly et al., 2015). For example, psychological distress can lead to self-medicating with alcohol (Phillips & Johnson, 2001; Turner, Mota, Bolton, & Sareen, 2018), comfort eating (Gibson, 2012), and smoking (Breslau, Novak, & Kessler, 2004), and it can be a motivational barrier to taking exercise (Firth et al., 2016).

Mental-health related differences in health behaviours may have widened during the pandemic reflecting the additional volitional efforts required to undertake such health behaviours during a lockdown; common mental health problems may lead to multiple barriers to undertaking such behaviours (e.g. feeling tired, loss of enjoyment in activities). Another explanation may be may a worsening of mental health symptoms (Henderson, 2020; Niedzwiedz et al., 2021), and thereby a worsening of health behaviours. Such worsening may be explained by muliple factors such as financial insecurity and changes to support mechanisms particularly affecting those with preeding mental health problems. Further research and examination will be needed to illuminate such mechanisms.

### Strengths and Limitations

Our study benefits from a large sample of participants from four UK cohort studies, spanning from age 19 to age 62. Because these cohorts have been followed longitudinally prior to the pandemic it was possible to examine previous measures of mental health and not just mental health concurrent with the pandemic. It is to our knowledge the first study to provide evidence on the effect of the pandemic on widening health behaviour inequalities based on mental health in the UK.

Whilst this study has many strengths it is important to carefully consider its limitations. As in many other COVID-19 surveys, fieldwork was planned and carried out rapidly. The online format used is likely to have contributed to the low response rates also observed in other comparable national studies (Niedzwiedz et al., 2021). While non-response weights (developed using individual and demographic data from previous sweeps) were used in analyses, we cannot fully exclude the possibility of there being unobserved predictors of missing data influencing our results. Other limitations relate to the self-reported the measures of health behaviours and mental health. In terms of health behaviours, although the recall period for pre-pandemic health behaviours was short, recall bias may have affected these measures. And it is possible that those with mental health problems may be especially affected by such recall bias, and respond differentially to health questions, potentially biasing associations.

Further, limited aspects of each health behaviours were used which do not include the full spectrum of these behaviours’ impact on health. For example, exercise does not capture less intensive physical activities, or sedentary behaviour; while fruit and vegetable intake is only one component of diet; and sleep is limited to sleep duration and not quality of sleep. In addition, the health behaviour risks may not be completely aligned with established health guidelines. Regarding mental health, prior and current psychological distress measures were not the same across cohorts, and the timing of their measurement prior to the pandemic varied across cohorts, meaning that any cohort differences could be due to a difference in measures and timing. Although the very similar results between different measures and the same measure of current mental health are encouraging and suggest little impact of how mental health is measured. As in all studies examining potential effects of the COVID-19 lockdown, we cannot distinguish whether differences found are due to different lockdowns or other time-varying factors such as seasonal change. Further, if such factors influenced mental health differentially this may account for changes in inequalities in health behaviour risks between timepoints.

### Conclusion

This study highlights the sizable inequalities in multiple health behaviours attributable to mental ill-health and shows how the COVID-19 lockdown may have further amplified these inequalities. This may have long-lasting effects on subsequent mental and physical health outcomes. The promotion of mental health may thus be an important component of improving post-COVID population health.

## Data Availability

The data used are available from the UK Data Archive

https://www.data-archive.ac.uk

## Other information

## Acknowledgements

We thank the Survey, Data, and Administrative teams at the Centre for Longitudinal Studies and Unit for Lifelong Health and Ageing, UCL, for enabling the rapid COVID-19 data collection to take place. We also thank Professors Rachel Cooper, Mark Hamer and Dr Jane Maddock for helpful discussions during the COVID-19 questionnaire design period.

## Financial support

PP and DB are supported in this work by funding from the UKRI via the Economic and Social Research Council (grant number ES/V012789/1). DB is supported by the Economic and Social Research Council (grant number ES/M001660/1) and Medical Research Council (MR/V002147/1). DB and AV are supported by The Academy of Medical Sciences / Wellcome Trust (“Springboard Health of the Public in 2040” award: HOP001/1025). PP is supported by the COVID-19 Longitudinal Health and Wellbeing National Core Study funded by the Medical Research Council (MC_PC_20030).

## Conflict of interest

All authors declare that they have no conflicts of interest with this manuscript.

## Ethical standards

The authors assert that all procedures contributing to this work comply with the ethical standards of the relevant national and institutional committees on human experimentation and with the Helsinki Declaration of 1975, as revised in 2008.

## Data availability

The data used are available from the UK Data Archive: https://www.data-archive.ac.uk.

## Notes

### Competing Interest Statement

The authors have declared no competing interest.

### Author Declarations

UCL/IOE ethics review board: https://www.ucl.ac.uk/ioe/research/research-ethics/ethics-applications-ioe-staff

### Summary of Updates

Title changed from five to four cohort studies. Also further funding sources were added.

