## Supplemental material for "Mental health in relation to changes in sleep, exercise, alcohol and diet during the COVID-19 pandemic: examination of four UK cohort studies" (from another version of this preprint: version 1, 2021-03-28)

**Figure S2. Differences in health behaviour risk (before COVID-19 pandemic, during May 2020 lockdown, and in September 2020) between participants with high and low levels of mental health problems: Examination of gender differeces.**
Note: Estimates show the risk difference on the percentage scale between those with the highest versus lowest mental health problems (ridit scores). Cohort and educational level are controlled for.

A: Psychological distress (prior to pandemic, cohort specific mental health measures)

**Before pandemic**

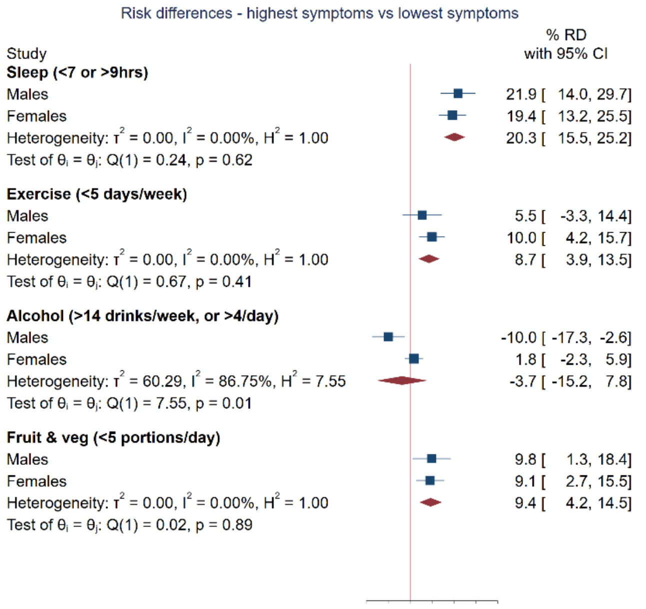


**May**

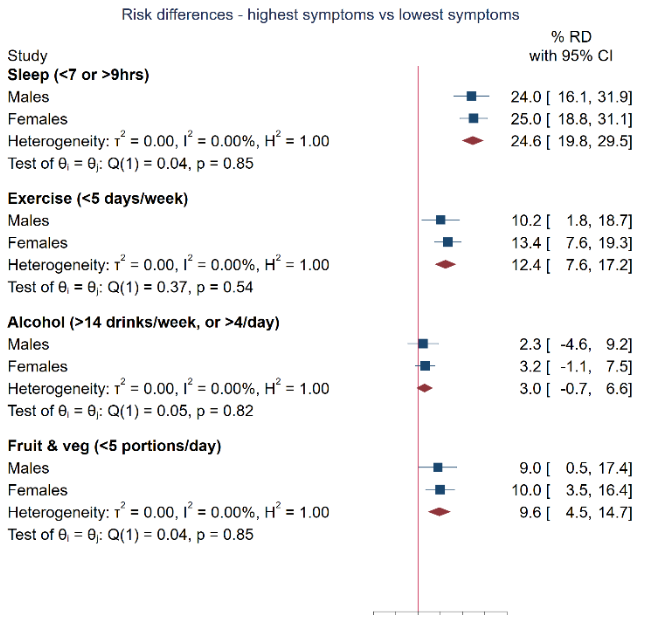


**September**

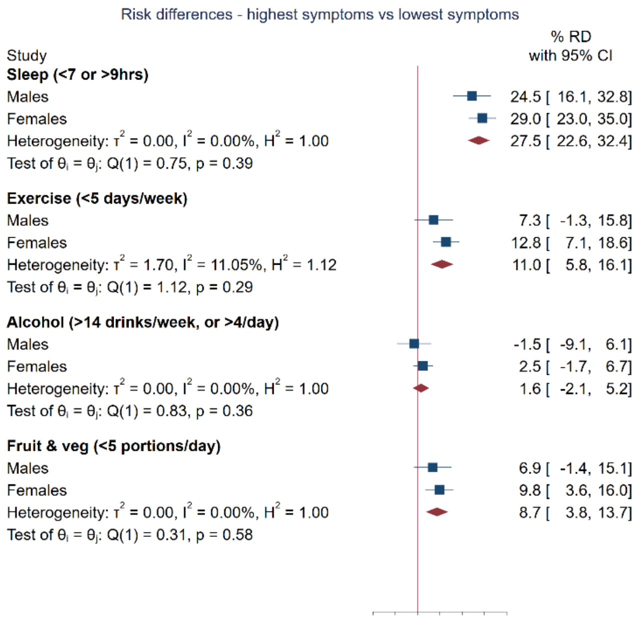

Figure S2 (continued)

B: Psychological distress (during May lockdown, cohort specific mental health measures)

### Before pandemic

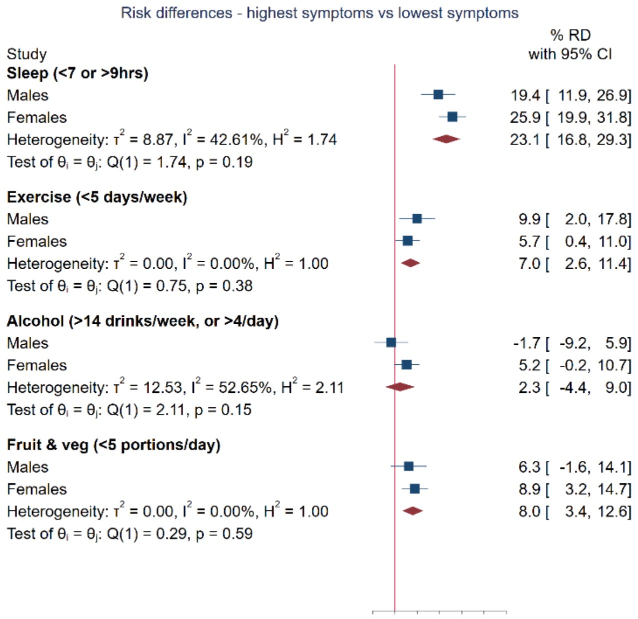


### May

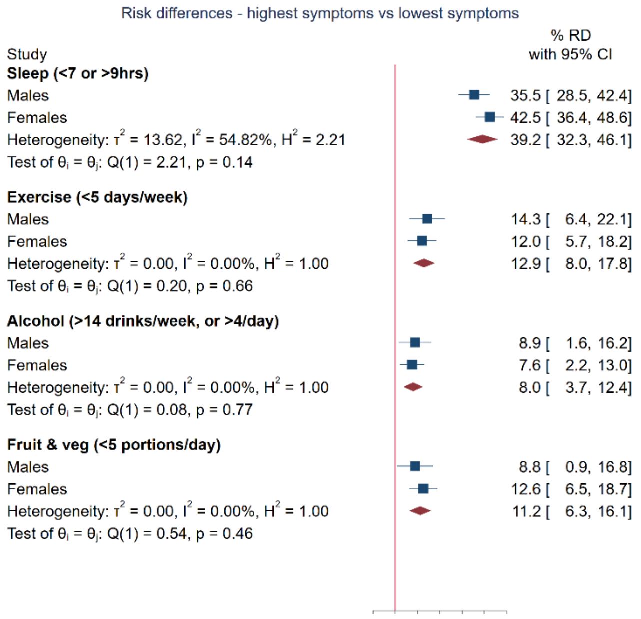


### September

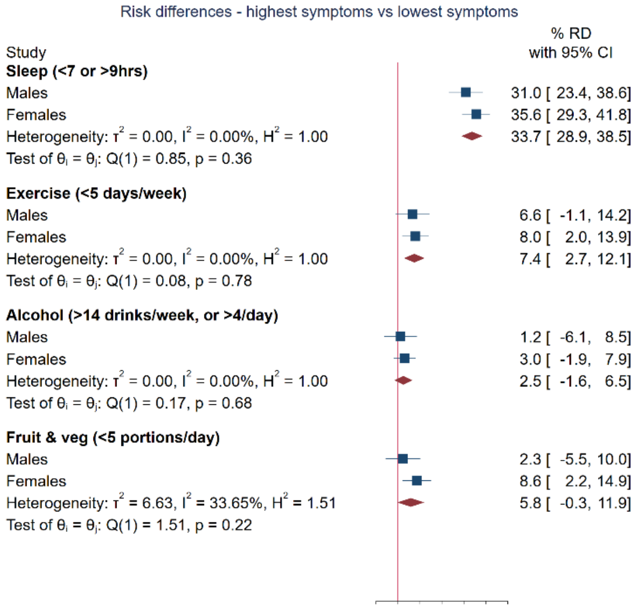

Figure S2 (continued)

C: Anxiety and depression (during May lockdown, same mental health measure across cohorts)

### Before pandemic

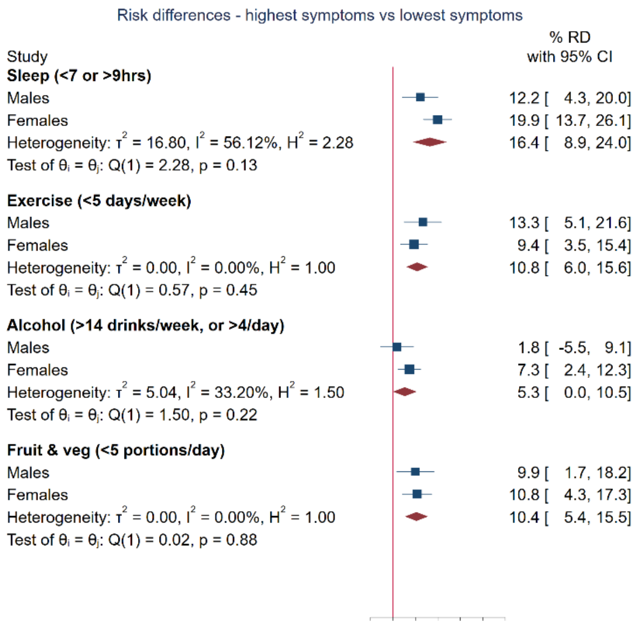


### May

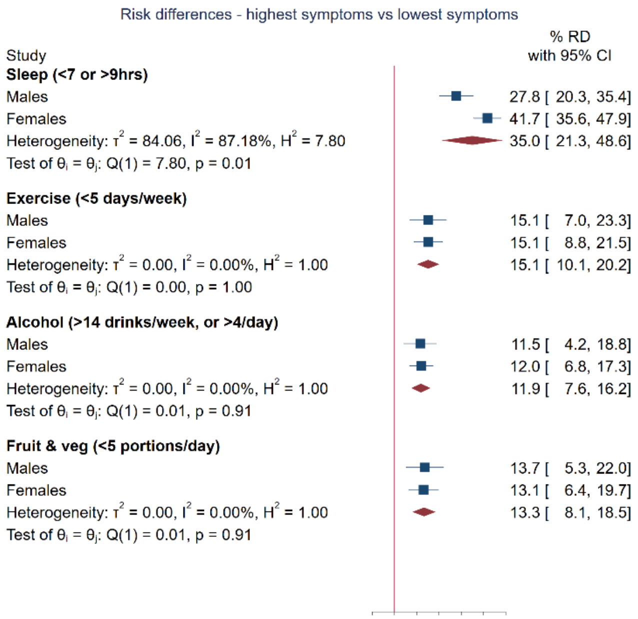


### September

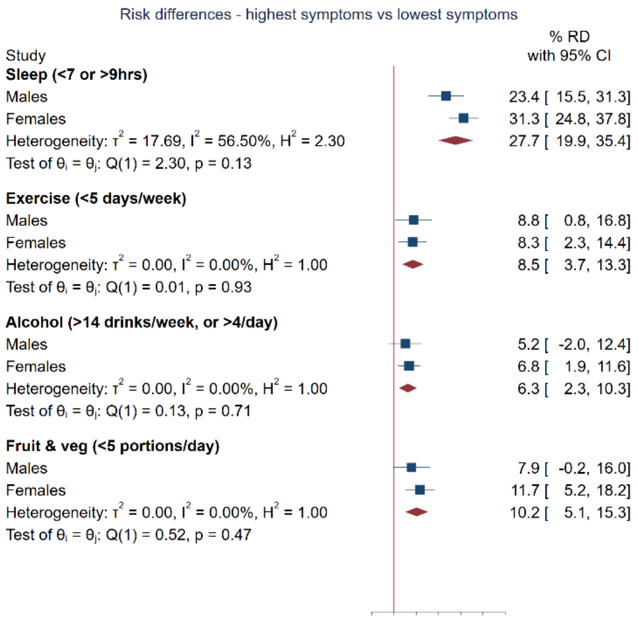

**Figure S3. Results of logistic regressions showing inequalities in health behaviour risk (before COVID-19 pandemic, during May 2020 lockdown, and in September 2020): meta-analysis of 5 cohort studies**

Note: Estimates show the risk difference on the percentage scale for each z-score difference in mental health symptoms, and are weighted to account for survey non-response and survey design in 2001c and 1990c. Sex and educational level are controlled for.

A: Psychological distress (prior to pandemic, cohort specific mental health measures)

**Before pandemic**

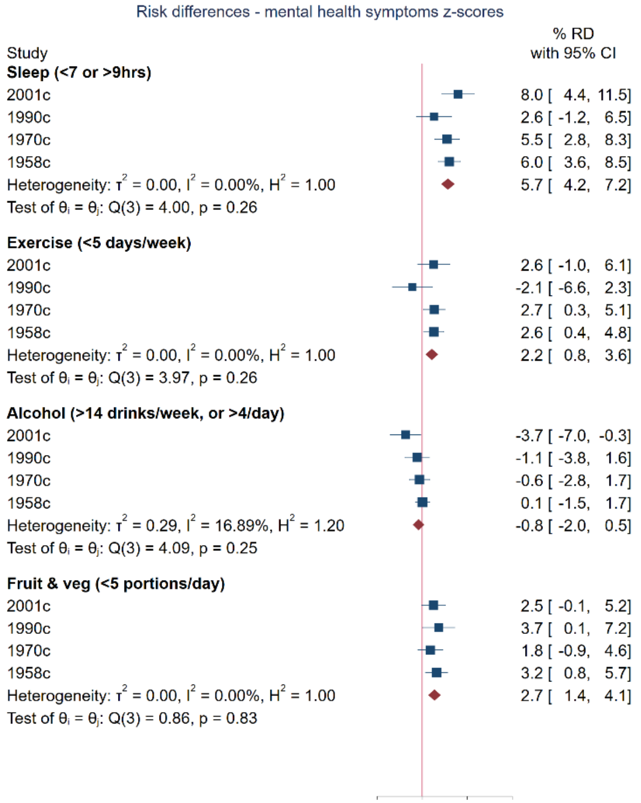


**May**

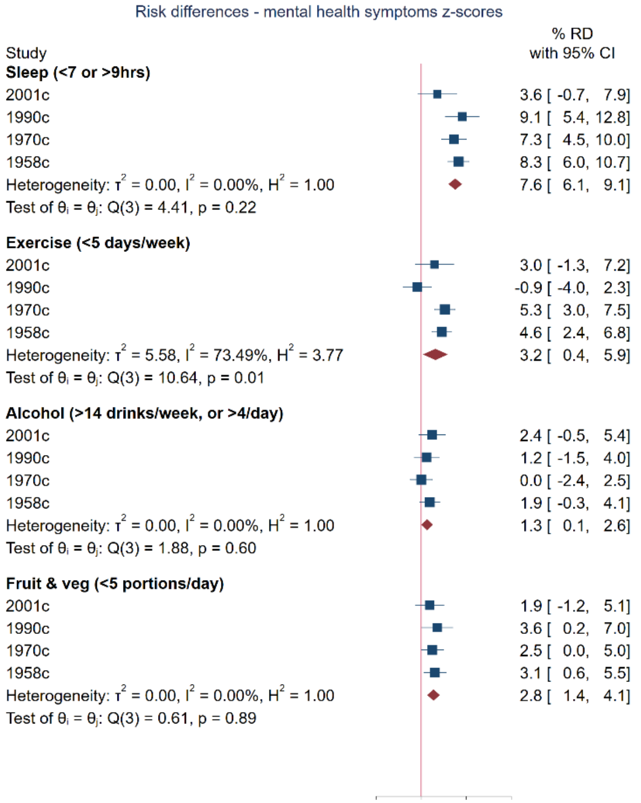


**September**

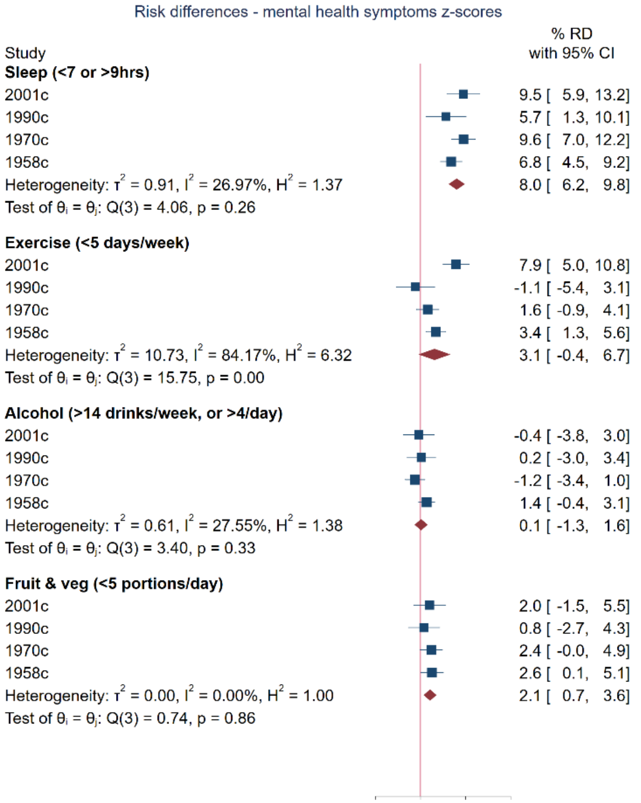

Figure S3: (continued)

B: Psychological distress (during May lockdown, cohort specific mental health measures)

**Before pandemic**

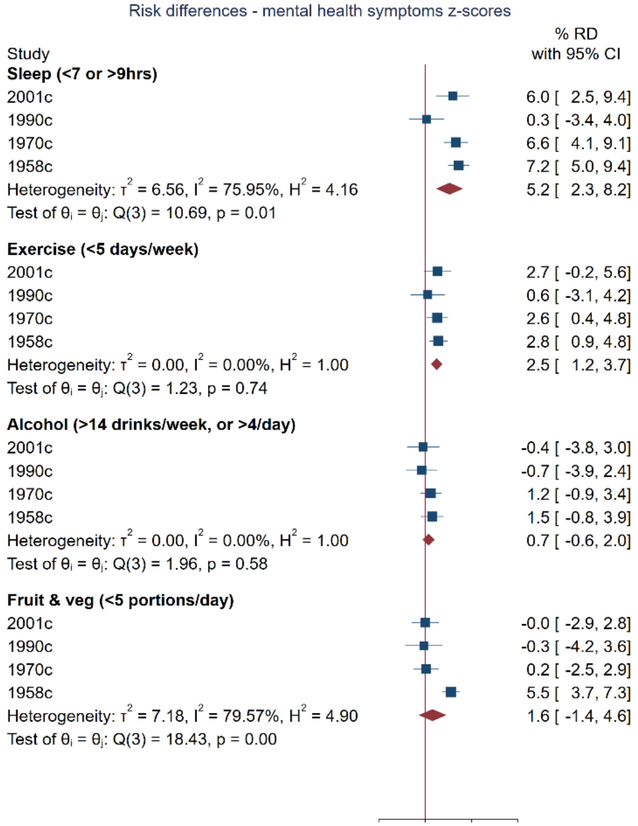


**May**

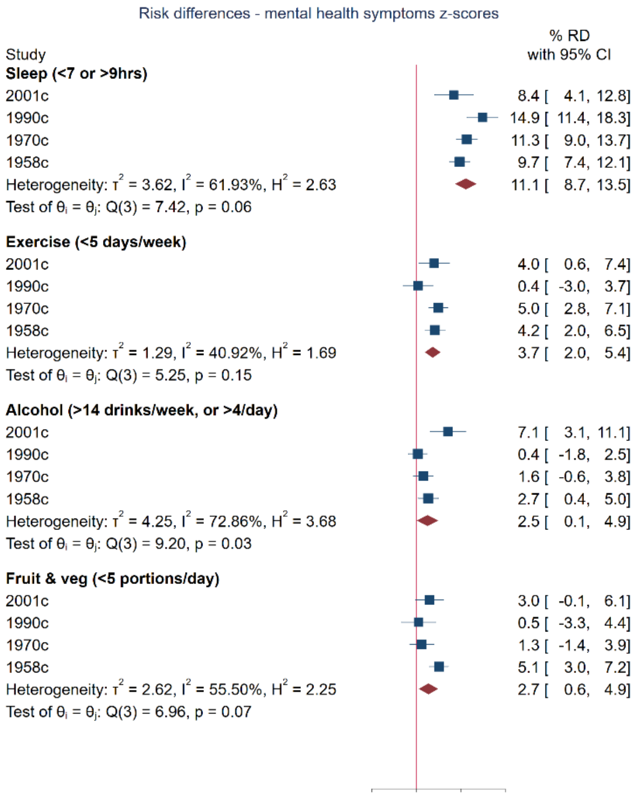


**September**

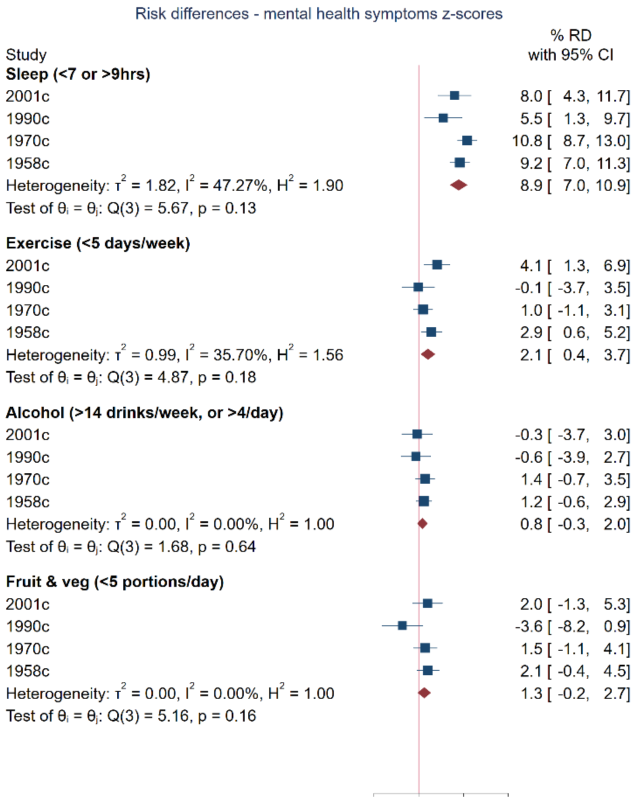

Figure S3: (continued)

C: Anxiety and depression (during May lockdown, same mental health measure across cohorts)

**Before pandemic**

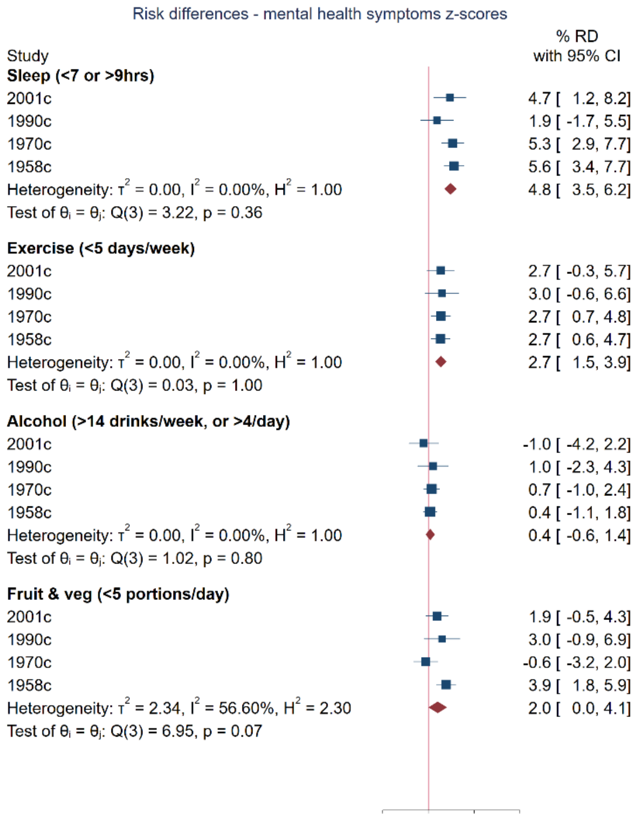


**May**

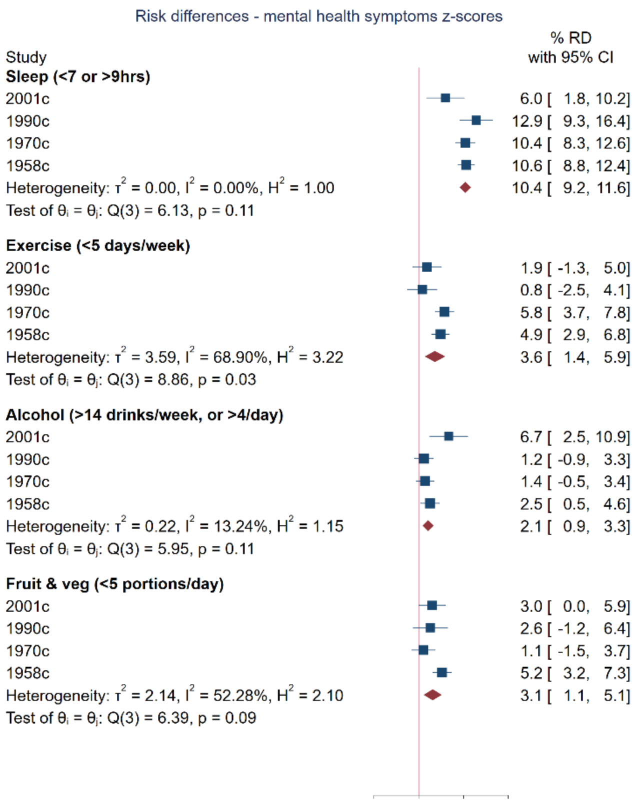


**September**

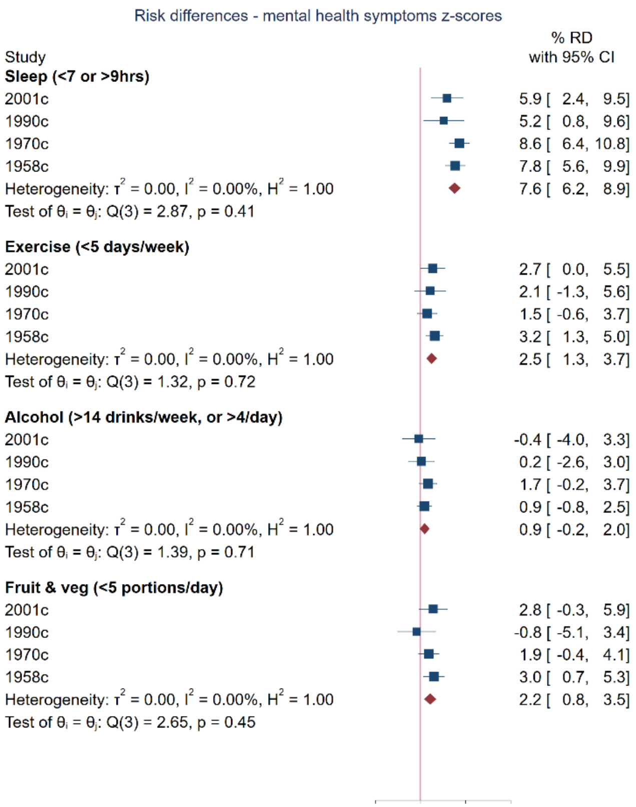

**Figure S4. Results of logistic regressions showing inequalities in health behaviour risk (before COVID-19 pandemic, during May 2020 lockdown, and in September 2020): meta-analysis of 5 cohort studies**

Note: Estimates show the risk difference on the percentage scale for those with clinical levels of mental health symptoms compared to non-clinical group, and are weighted to account for survey non-response and survey design in 2001c and 1990c. Sex and educational level are controlled for.

A: Psychological distress (prior to pandemic, cohort specific mental health measures)

**Before pandemic**

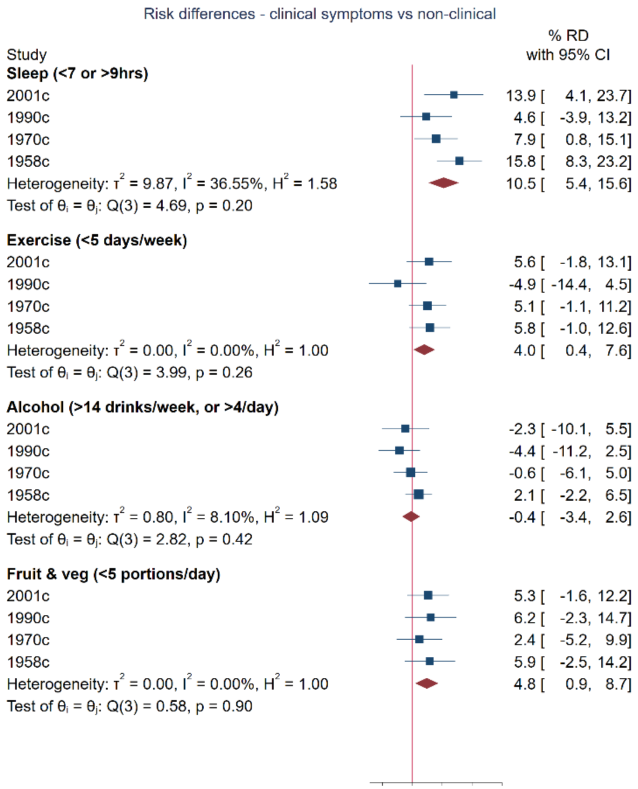


**May**

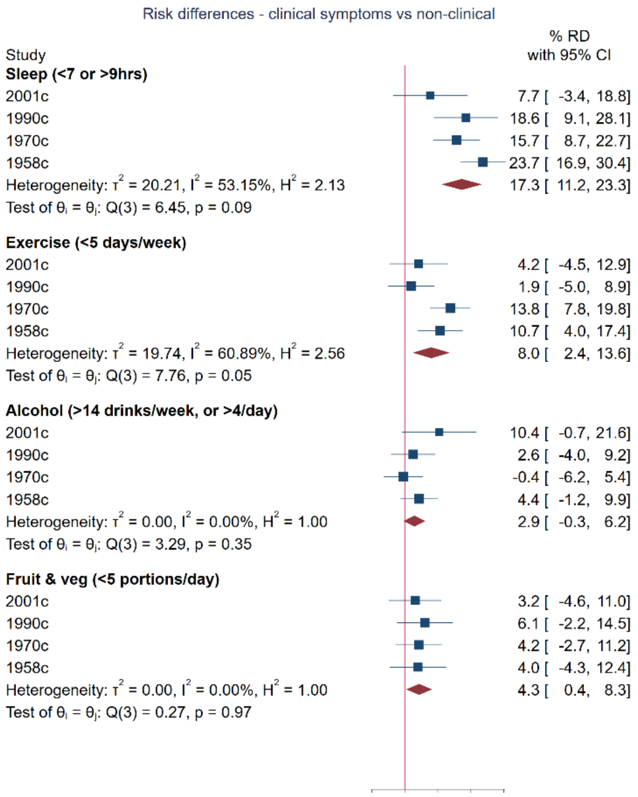


**September**

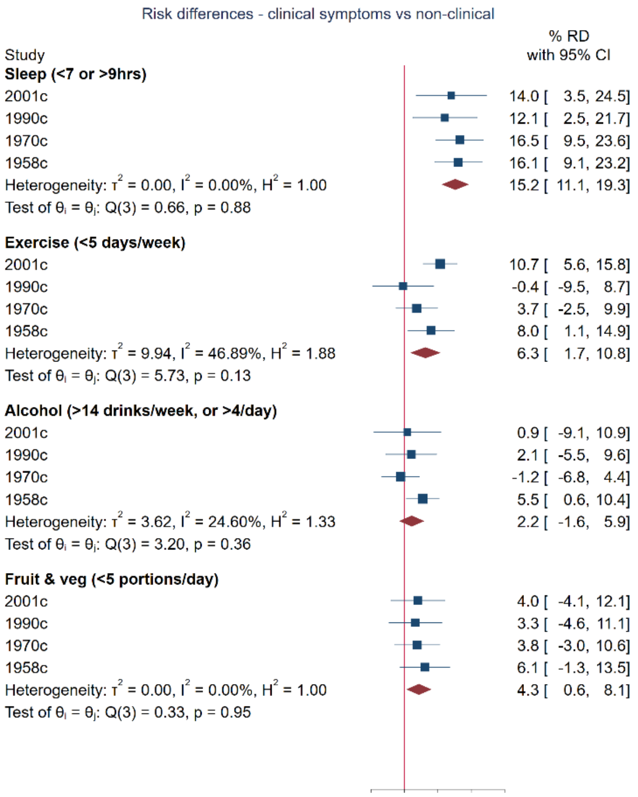

Figure S4: (continued)

B: Psychological distress (during May lockdown, cohort specific mental health measures)

**Before pandemic**

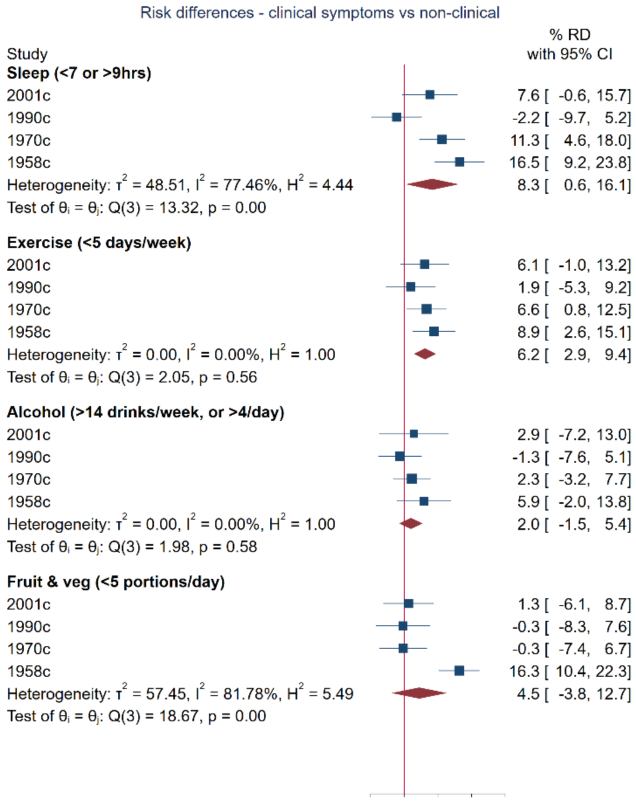


**May**

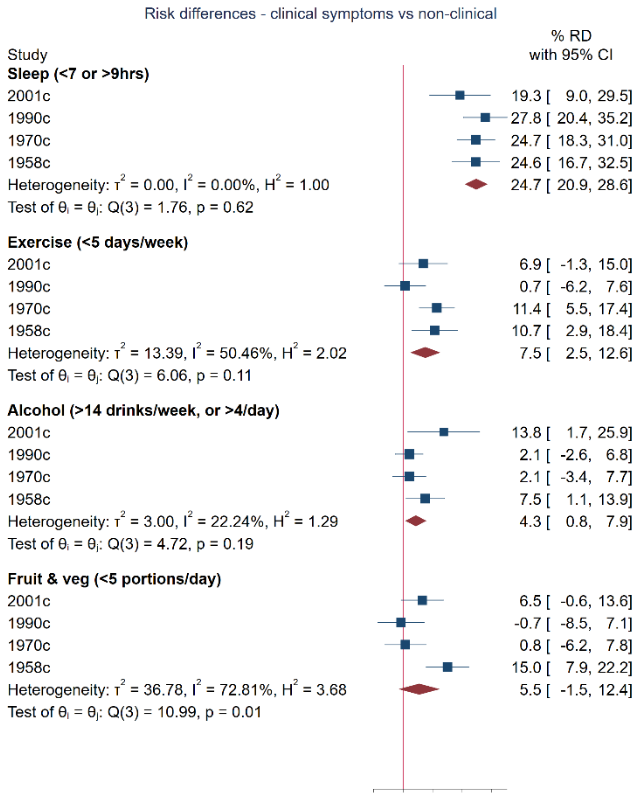


**September**

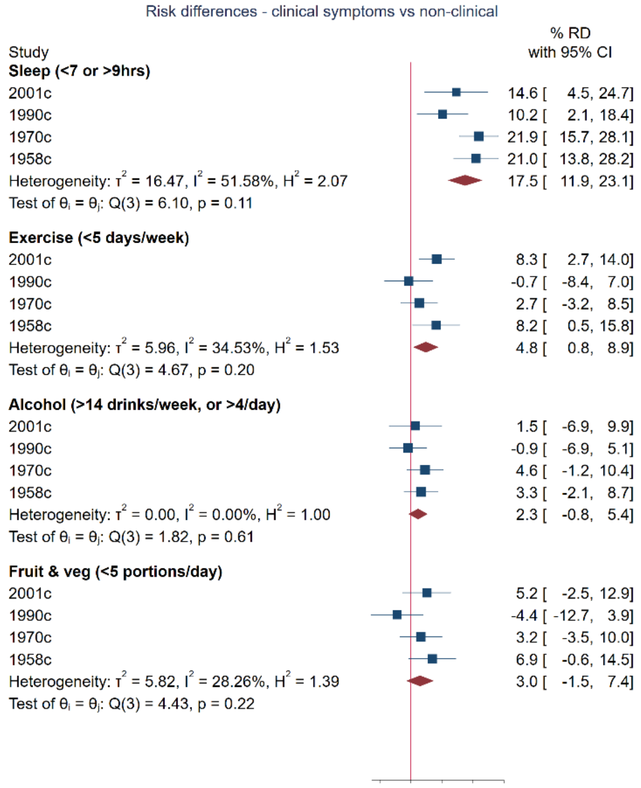

Figure S4: (continued)

C: Anxiety and depression (during May lockdown, same mental health measure across cohorts)

### Before pandemic

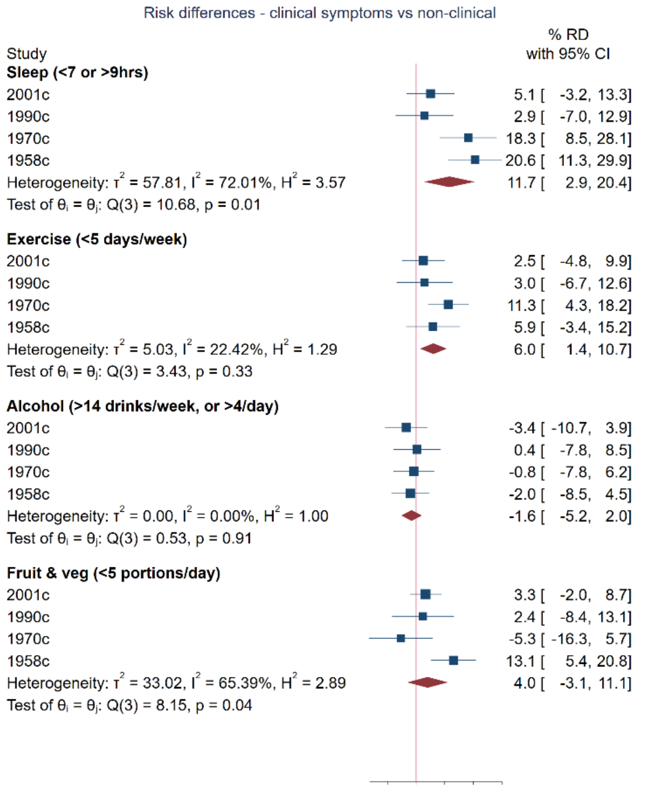


### May

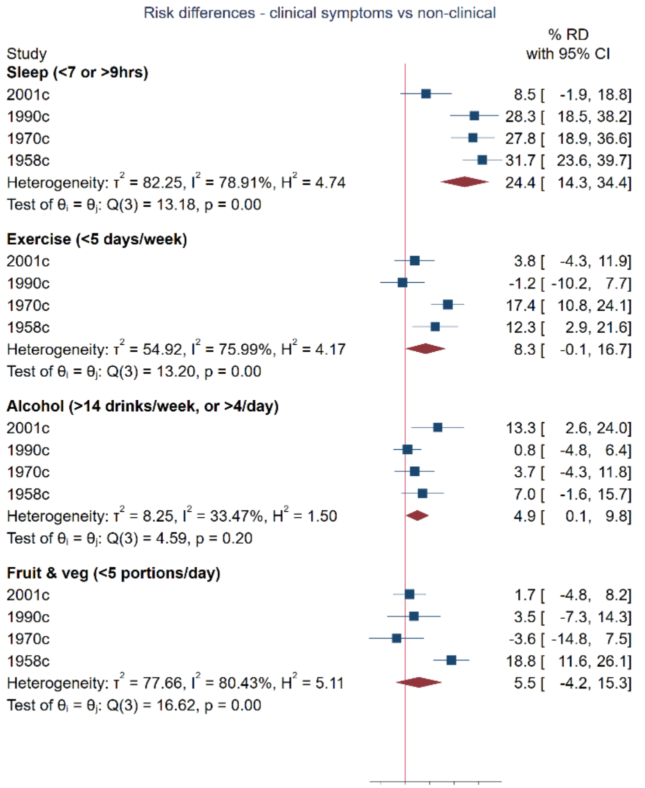


### September

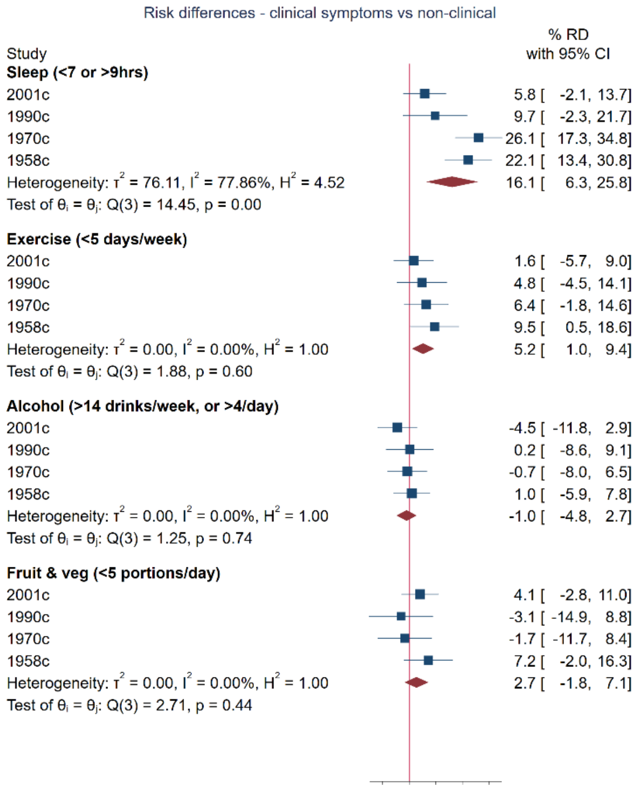
